# “Association between Median Household Income, State Medicaid Expansion Status, and COVID-19 Outcomes Across US Counties”

**DOI:** 10.1101/2021.02.03.21251075

**Authors:** Tsikata Apenyo, Antonio Vera-Urbina, Khansa Ahmad, Tracey H. Taveira, Wen-Chih Wu

**Author notes:** **Address for correspondence:** Wen-Chih Wu, MD., Providence VA Medical Center, Miriam Hospital Cardiovascular Rehabilitation Center, Alpert Medical School and School of Public Health at Brown University, Division of Cardiology, 830 Chalkstone Avenue, Providence, RI 02908.

## Abstract

**Objective:** The relationship between socioeconomic status and its interaction with State’s Medicaid-expansion policies on COVID-19 outcomes across United States (US) counties are uncertain. To determine the association between median-household-income and its interaction with State Medicaid-expansion status on COVID-19 incidence and mortality in US counties

**Methods:** Longitudinal, retrospective analysis of 3142 US counties (including District of Columbia) to study the relationship between County-level median-household-income (defined by US Census Bureau’s Small-Area-Income-and-Poverty-Estimates) and COVID-19 incidence and mortality per 100000 of the population in US counties from January 20, 2020 through December 6, 2020. County median-household-income was log-transformed and stratified by quartiles. Medicaid-expansion status was defined by US State’s Medicaid-expansion adoption as of first reported US COVID-19 infection, January 20, 2020. Multilevel mixed-effects generalized-linear-model with negative binomial distribution and log link function compared quartiles of median-household-income and COVID-19 incidence and mortality, reported as incidence-risk-ratio (IRR) and mortality-risk-ratio (MRR), respectively. Models adjusted for county socio-demographic and comorbidity conditions, population density, and hospitals, with a random intercept for states. Multiplicative interaction tested for Medicaid-expansion*income quartiles on COVID-19 incidence and mortality.

**Results:** There was no significant difference in COVID-19 incidence across counties by income quartiles or by Medicaid expansion status. Conversely, significant differences exist between COVID-19 mortality by income quartiles and by Medicaid expansion status. The association between income quartiles and COVID-19 mortality was significant only in counties from non-Medicaid-expansion states but not significant in counties from Medicaid-expansion states (P<0.01 for interaction). For non-Medicaid-expansion states, counties in the lowest income quartile had a 41% increase in COVID-19 mortality compared to counties in the highest income quartile (MRR 1.41, 95% CI: 1.25-1.59).

**Conclusions and Relevance:** Median-household-income was not related to COVID-19 incidence but negatively related to COVID-19 mortality in US counties of states without Medicaid-expansion. It was unrelated to COVID-19 mortality in counties of states that adopted Medicaid-expansion. These findings suggest that expanded healthcare coverage should be investigated further to attenuate the excessive COVID-19 mortality risk associated with low-income communities.

**Key Findings:** *Question:* Is there a relationship between COVID-19 outcomes (incidence and mortality) and household income and status of Medicaid expansion of US counties?

*Findings:* In this longitudinal, retrospective analysis of 3142 US counties, we found no significant difference in COVID-19 incidence across US counties by quartiles of household income. However, counties with lower median household income had a higher risk of COVID-19 mortality, but only in non-Medicaid expansion states. This relationship was not significant in Medicaid expansion states.

*Meaning:* Expanded healthcare coverage through Medicaid expansion should be investigated as an avenue to attenuate the excessive COVID-19 mortality risk associated with low-income communities.

## Background

Instituted in the 1965, Medicaid has become the largest provider of health insurance in the United States (US) by providing medical access to people with low income and limited resources.^1^ The 2010 Affordable Care Act (ACA) sought to decrease the number of uninsured individuals by expanding Medicaid coverage and modifying individual insurance markets,^2,3,4^ but a 2012 Supreme Court decision overturned the requirement that states adopt Medicaid expansion.^4,5^ By January 2020, 14 states had yet to adopt Medicaid expansion.^6^ Studies have found that among states who implemented the ACA, the increased access to care has led to early diagnosis of cancers, diabetes, and depression among other health outcomes,^7-10^ but the relationship between Medicaid expansion and an infectious disease outbreak is unknown.

In 2019, a novel coronavirus (COVID-19), originating from Wuhan City, Hubei Province, China began spreading at an alarming rate.^11^ As COVID-19 progressed in the United States (US), the health toll disproportionally impacted African Americans and communities with high prevalence of poor housing conditions.^12-14^ In addition, COVID-19 has already been shown to impact individuals with certain pre-existing health conditions at a greater rate.^15^ Both federal and state policymakers looked to Medicaid as a central tool in their response to the national emergency.^16^ However, whether differences exist in COVID-19 outcomes between communities of Medicaid and non-Medicaid expansion states remains unknown. Moreover, it would be important to quantify differences in outcomes, if any, on the strata that appeared most impacted by COVID-19, the low-income communities, to help the states balance cost versus benefits. This is important because individuals without health insurance coverage are likely to be more vulnerable to the adverse health outcomes related to COVID-19. In the states that did not implement Medicaid expansion, 30 percent of low-income workers were uninsured before COVID-19.^17^ This number was less than half in Medicaid expansion states.^17^

We thereby sought to investigate the impact of COVID-19 in the counties nationwide according to their socio-economic status and investigate whether the impact varies by counties of states with Medicaid expansion versus those without Medicaid expansion. We stratified the US counties by its median household income and compared on their COVID-19 incident and mortality rates. We hypothesized that household income would be inversely related to COVID-19 incidence and mortality. Additionally, we hypothesize that a state’s Medicaid expansion status will alter the association between median household income and COVID-19 outcomes.

## Methods

We conducted a longitudinal, retrospective analysis of data of the US counties and District of Columbia (n = 3142) using 2010-2019 baseline data from the Centers for Disease Control and Prevention and the US Census Bureau and related them to the COVID-19 outcome data from the John Hopkins Coronavirus Resource Center, 2020. ^18-21^ Counties from US territories (American Samoa, Guam, Northern Mariana Island, Puerto Rico and US Virgin Islands, n=105) were not part of the analysis.^18^ All data used in this study were publicly available; therefore, the study met the criteria for exemption by the Providence Veterans Affairs Medical Center Institutional Review Board.

### Main Exposure Variables

Median household income for each county was collected from the 2018 US Census Bureau’s Small Area Income and Poverty Estimates (SAIPE),^19^ and log-transformed (ln X +1) to approximate normality prior to stratification by quartiles. We defined Medicaid expansion states as those that had adopted expansion efforts as of the first case of COVID-19 in the United States on January 20, 2020 (Listing in Supplement).^6^ Counties in 36 states plus Washington, DC, were included in the Medicaid expansion group, while counties in 14 states were in the non-Medicaid expansion group.

### Outcome

The main outcomes of our study were COVID-19 incidence and mortality per 100000 of the population. The absolute COVID-19 incidence and mortality rates of the US counties were obtained from the John Hopkins Coronavirus Resource Center and divided by the county population. Data from January 20, 2020 until December 6, 2020 were utilized for the analyses.^21^

### Covariates

Data on age and gender were collected using 2010 US Census Bureau data as the elderly and men have been reported as possessing a higher risk of COVID-19 mortality.^19^ The COVID-19 pandemic has been shown to afflict minority races in the US to a greater degree; therefore, we included data for racial composition of counties: percentage of White, Black, and Hispanic residents using US Census Bureau data from 2014-2018. Population density (population per square feet of land area) was calculated from the county population from 2010 US census divided by the square foot are of the county to account for overcrowding in a community. In addition to median-household income, we also abstracted data that were confirmatory of the socioeconomic status of the communities such as unemployment rate (2019), percentage of population age > 25 years without high school diploma (2014-2018), and percentage of population age < 65 years without health insurance (2018).^22^ Access to care was assessed by number of hospitals per county (2017).

Since diabetes mellitus, obesity, and smoking are known risk factors for worse outcomes in COVID-19, ^23^ we obtained the percentage of the population aged >20 years diagnosed with diabetes mellitus, with obesity, and percentage of adults who are current smokers from the Centers for Disease Control and Prevention from 2016-2018.^20,23^

### Statistical Analysis

Baseline characteristics for the counties were described by mean ± standard deviation (SD) and range for continuous variables and percentage for categorical variables. Counties were stratified by quartiles of log-transformed median household income, which comprised of the following median household income ranges: Q1 ($25385 - $43681); Q2 ($43688 - $50565); Q3 ($50568 - $58838); Q4 ($58848 - $140382). Linear regression was used to test for trend of baseline characteristics across the income quartiles.

We used a multilevel mixed-effects generalized linear model with a negative binomial distribution and log link function to study the relationship between quartiles of log-median household income and COVID-19 outcomes across US counties: incidence and mortality, in a separate fashion, using Q4 as the referent. We applied a random intercept for states to account for clustering effect due to similarities in health policy for counties within the same state and specifying an unstructured covariance matrix. Using county population as the denominator in the model, the outcomes reported were incidence rate ratios (IRR) and mortality rate ratios (MRR) of COVID-19 across income quartiles of the counties, respectively. In a stepwise fashion, we first adjusted for demographics age over 65 years old, gender, and race (Model 1); followed by population density, diabetes, obesity, current smoking status, state Medicaid expansion status and number of hospitals (Model 2). The percentage of population without high school diploma under 25 years old and population without health insurance were not included in the model given their significant correlation with the median household income per county (*r*= −0.36, *P*<0.001) and the Medicaid expansion status (*r*= −0.63, *P<*0.001) variables, respectively. We tested for interaction between quartiles of log-median household ‘income quartile*Medicaid expansion status’ on COVID-19 outcomes in Model 2. If the interaction proved to be significant, the above analyses were repeated stratified by counties of states with Medicaid Expansion (n=1814) and counties of states without Medicaid Expansion (n=1328).

Sensitivity analyses were performed to replace median age in lieu of % over 65 years old. All analyses were performed using STATA/SE version 11.2 software (StataCorp LP, College Station, TX). A 2-sided p-value of < 0.05 was considered significant.

## Results

As of December 6, 2020, there were a total of 14528356 COVID-19 cases, and 279115 COVID-19 deaths, across the 3142 counties. The mean (SD) for COVID-19 incidence and mortality were 5155.01 (4308.24) cases and 88.38 (96.46) deaths per 100000 population per county, respectively.

The characteristics of the 3142 counties, overall and stratified by four quartiles of log-transformed median household income are presented in Table 1. Overall, 57.7% of counties were located in Medicaid expansion states. Higher median household income quartiles were associated with higher mean county population and population density, number of hospitals and percentage of white residents. Conversely, lower median household income quartiles were associated with higher percentage of elderly residents (65 years or older), black or Hispanic, unemployed and who did not have high school diploma or health insurance. Counties of lower income quartiles were also associated with a higher prevalence of diabetes, obesity and smoking and had a lower likelihood of belonging to a state that adopted Medicaid-expansion.

**Table 1:**
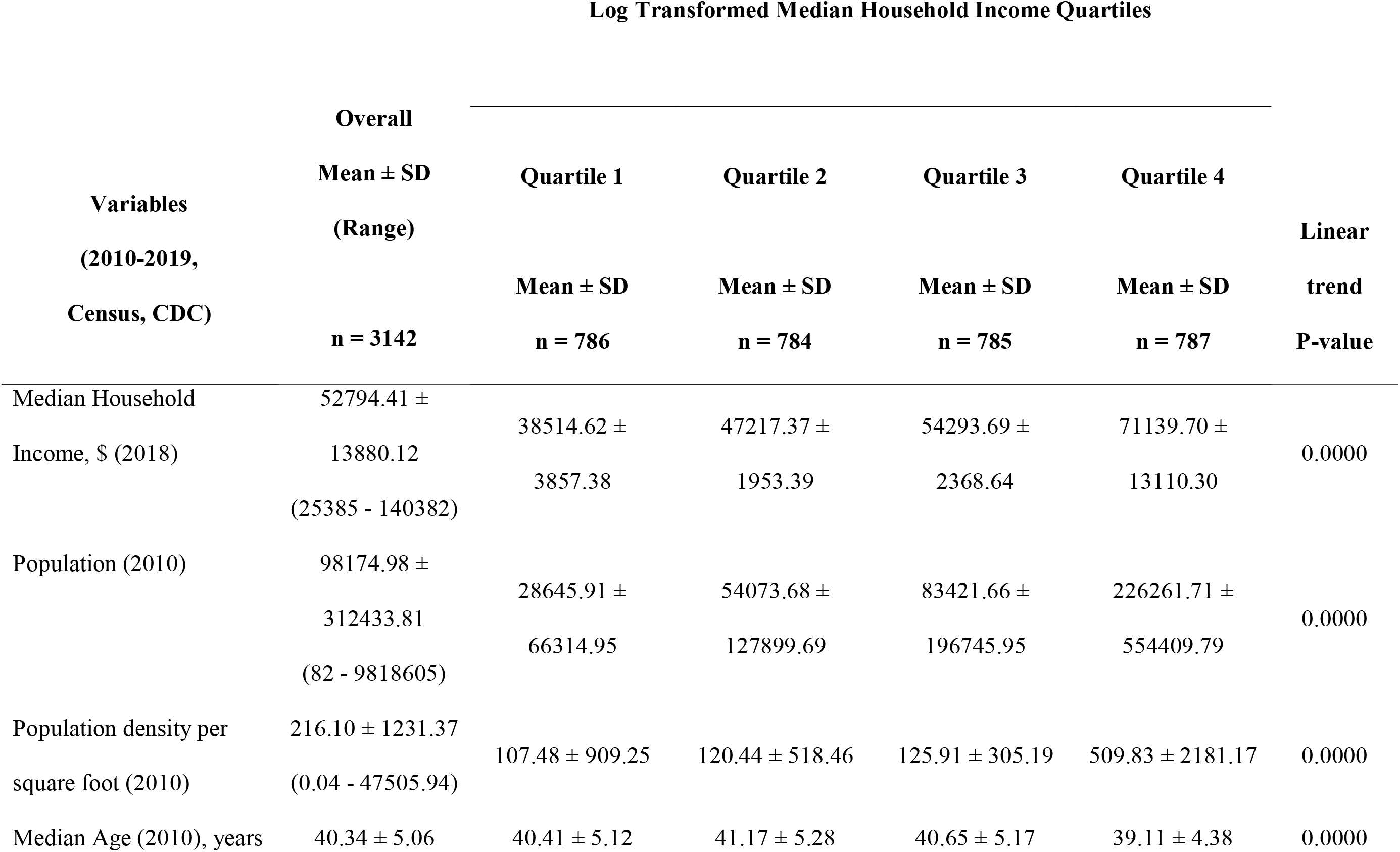
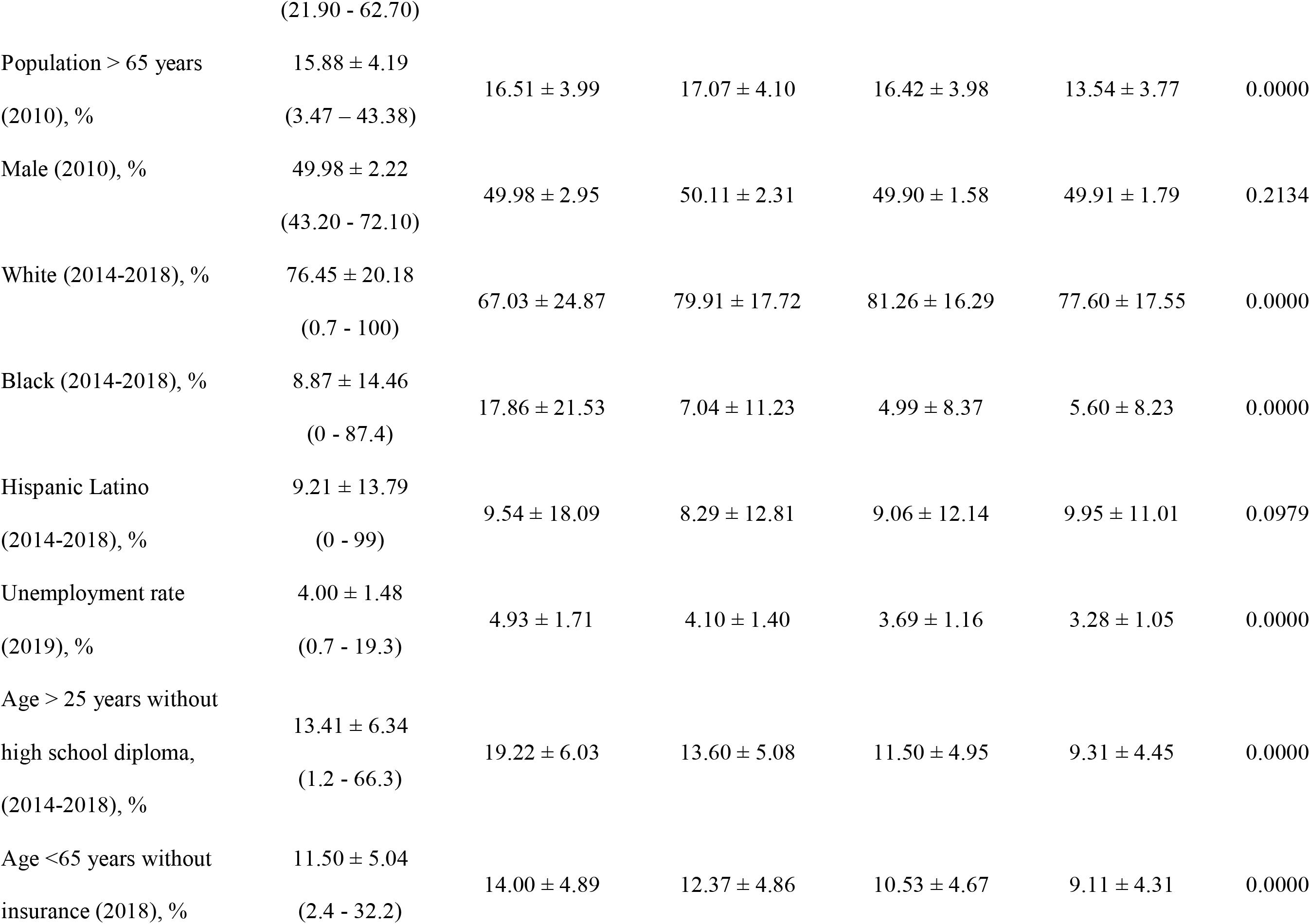
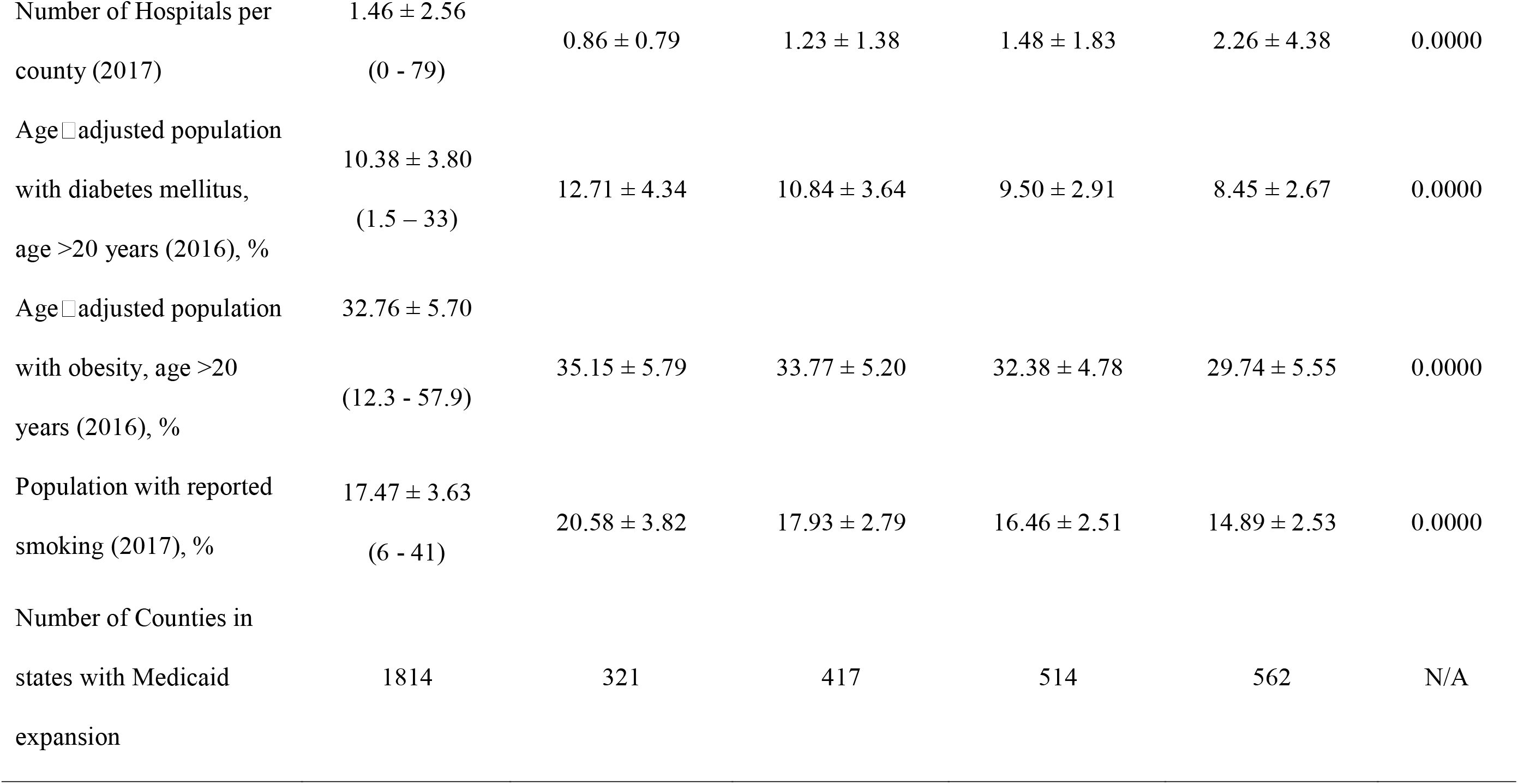
Counties Baseline Characteristics by Log Transformed Median Household Income

The mean number of COVID-19 cases and deaths per 100000 population across counties from different income quartiles were described in Table 2. There was no significant association between COVID-19 incidence and quartiles of household income in unadjusted and demographic adjusted analyses. In the fully adjusted model, counties from income quartile 2 had a 10% increase in the risk of COVID-19 incidence compared to quartile 4 (IRR 1.10, 95% CI: 1.04-1.17). The interaction ‘income quartile*Medicaid expansion status’ was not significant for COVID-19 incidence (P values 0.07 to 0.20 Q1-3).

**Table 2:** Association of SARS-COV-2 outcomes as of December 6, 2020 with Median Household Income Quartiles

| SARS-CoV2 Outcomes<br>(As of December 6, 2020) | Median Household Income Quartiles |  |  |  |
| --- | --- | --- | --- | --- |
|  | **Quartile 1** | **Quartile 2** | **Quartile 3** | **Quartile 4** |
|  | N= 786 | N= 784 | N= 785 | N= 787 |
|  | IRR / MRR<br>(95% CI) | IRR / MRR<br>(95% CI) | IRR / MRR<br>(95% CI) | IRR / MRR<br>(95% CI) |
| Cases per 100,000 population<br>(mean ± SD) | 5121.08 ± 2471.59 | 5299.32 ± 5338.71 | 5166.40 ± 2666.65 | 5033.77 ± 5705.18 |
| Model 1 | 0.96<br>[0.90-1.02] | 1.05<br>[0.99-1.10] | 0.96<br>[0.91-1.01] | REFERENT |
| Model 2* | 1.04<br>[0.97-1.12] | 1.10<br>[1.04-1.17] | 1.00<br>[0.95-1.05] | REFERENT |
| Deaths per 100,000 population<sup>#</sup><br>(mean ± SD) | 113.32 ± 87.43 | 92.21 ± 109.58 | 75.69 ± 62.84 | 72.32 ± 112.19 |
| Model 1 | 1.16<br>[1.06-1.26] | 1.15<br>[1.07-1.24] | 0.99<br>[0.93-1.06] | REFERENT |
| Model 2* | 1.22<br>[1.09-1.35] | 1.18<br>[1.08-1.28] | 1.02<br>[0.95-1.10] | REFERENT |

IRR = Incident Rate Ratio; MRR = Mortality Rate Ratio; 95% CI = 95% Confidence Interval
Model 1: % Population > 65 years, % Male, and % White
Model 2: % Population > 65 years, % Male, % White, Population Density, % Obesity, % Smoking, % Diabetes, Number of Hospitals, Medicaid expansion status according to state policy
\*Interaction between income quartiles and Medicaid status was significant ( $p\text{-value} \leq 0.005$ ) for SARS-COV-2 mortality but not for SARS-CoV2 Cases ( $p\text{-value} \geq 0.073$ )

Conversely, there was a significant association between COVID-19 mortality and quartiles of household income. In the fully adjusted model, counties from income quartile 1 had a 22% increase in the risk of COVID-19 mortality compared to quartile 4 (MRR 1.22, 95% CI 1.09-1.35). Furthermore, the interaction ‘income quartile*Medicaid expansion status’ was significant (P values <0.01, Q1-3), for which subgroup analyses by Medicaid expansion status were conducted. The sensitivity analyses replacing % population over 65 years old with median age of the county population did not significantly change the results.

The comparison of baseline characteristics between counties in Medicaid and non-Medicaid expansion states were described in Table 3. Counties from states with Medicaid expansion had a higher population density, percentage of white residents, median household income, unemployment rate, number of hospitals; and a lower percentage of population who were Black, Hispanic, without high school diploma, without health insurance, with diabetes, with obesity or reported being a current smoker. The association between household income quartiles and COVID-19 mortality by state Medicaid expansion status was depicted in Figure 1 and described in supplemental Table 2. In fully adjusted analyses, median household income quartiles were associated with COVID-19 mortality only in counties within states without adoption of Medicaid expansion, such that counties in the lowest income quartile had a 41% increase in COVID-19 mortality compared to counties in the highest income quartile (MRR 1.41, 95% CI: 1.25-1.59). Contrarily, income quartiles were not associated with COVID-19 mortality in counties within states that adopted Medicaid expansion.

**Table 3:**
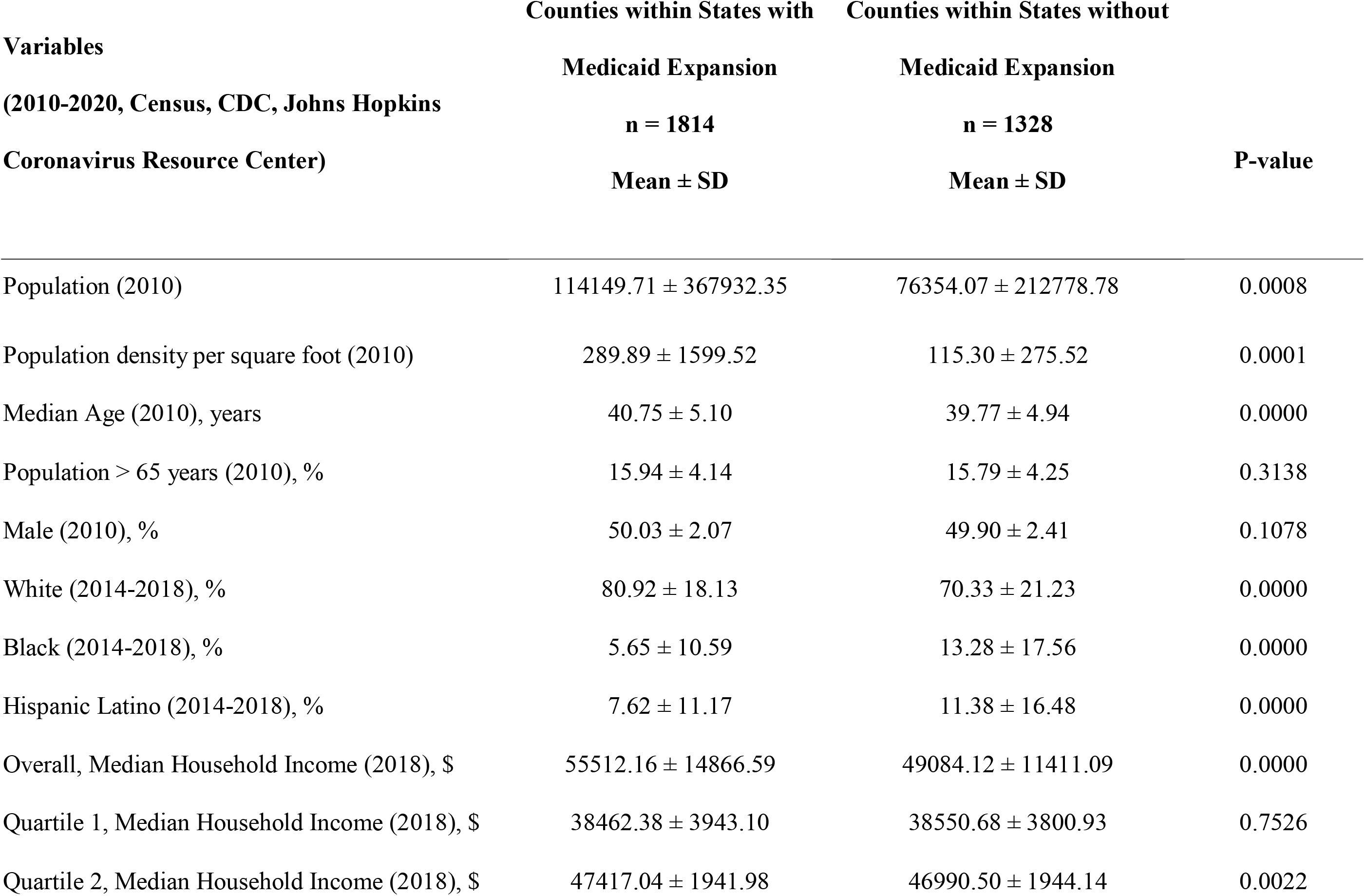
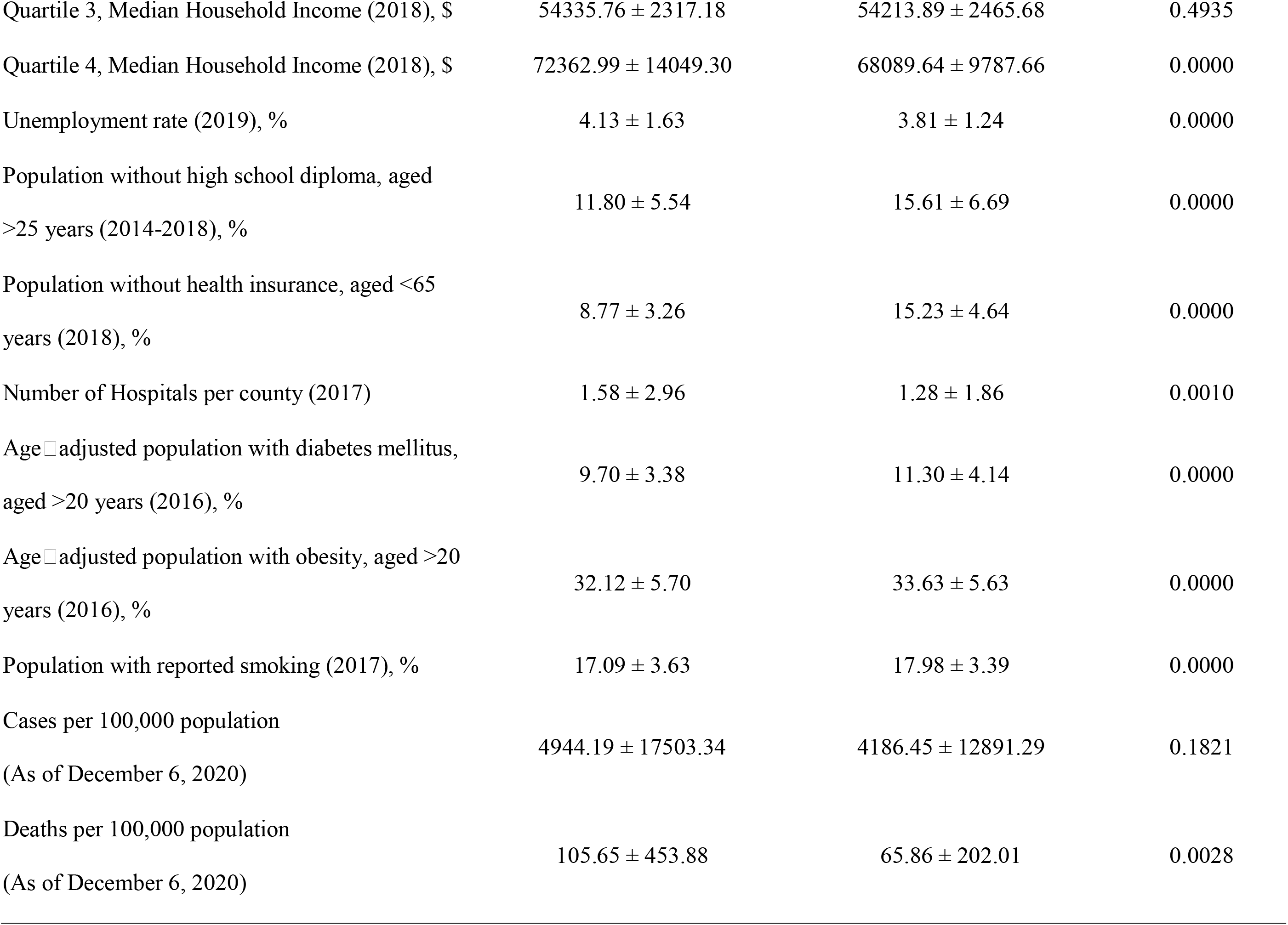
Counties Baseline Characteristics by Medicaid Expansion Status of the State

**Figure 1.**
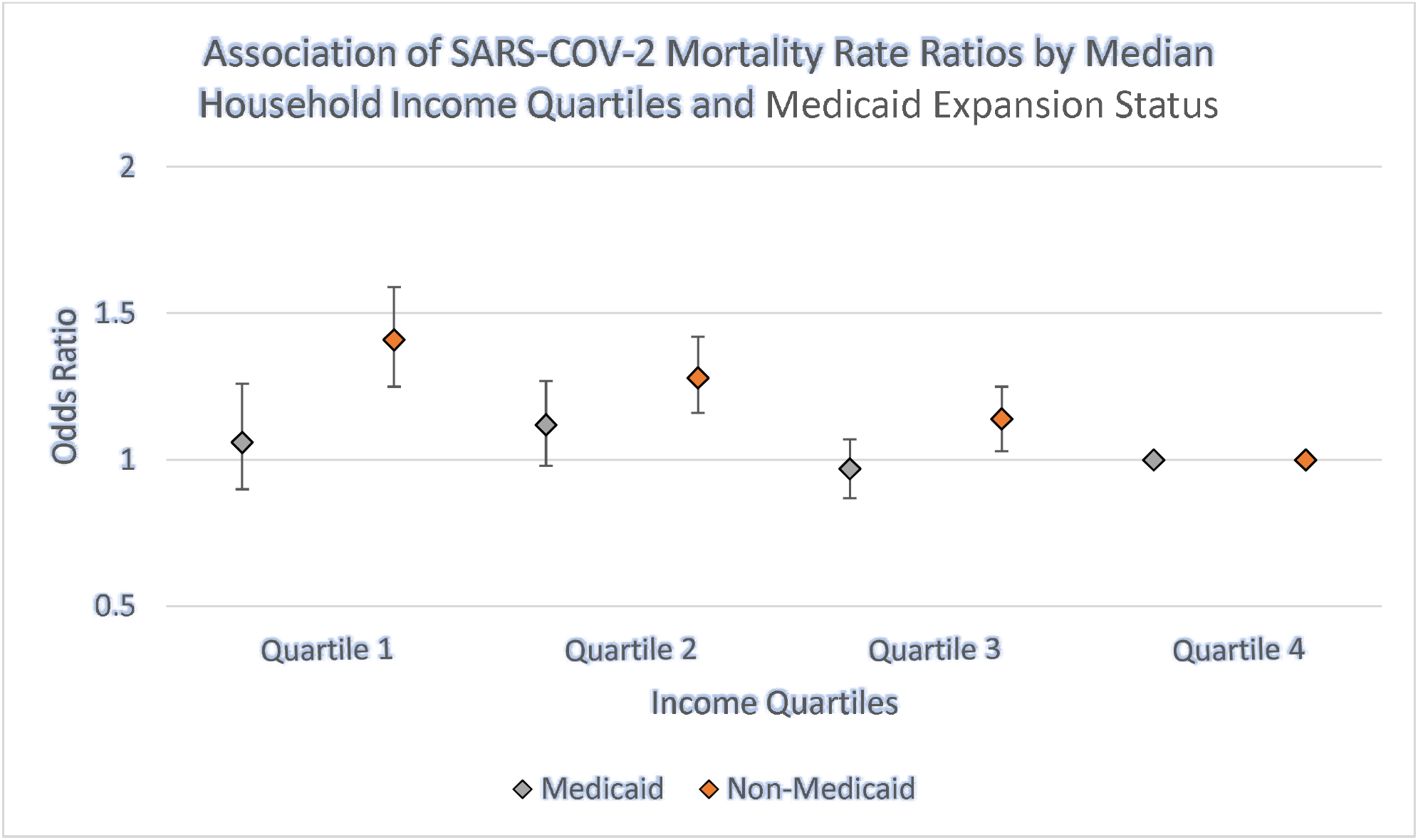
Association of SARS-COV-2 Mortality Rate Ratios by Median Household Income Quartiles, for counties in Medicaid and Non-Medicaid expansion states, referent = Quartile 4

## Discussion

To our knowledge, this is the first paper to study the association of median household income with COVID-19 outcomes at the county level, in Medicaid expansion and non-expansion states. We found no significant difference in COVID-19 incidence across counties by income quartiles or by Medicaid-expansion status. However, significant differences exist between COVID-19 mortality by income quartiles and by Medicaid-expansion status, such that the association between income quartiles and COVID-19 mortality was significant only in counties from non-Medicaid-expansion states but not significant in counties from Medicaid-expansion states.

There is ample evidence to support that socioeconomic status is related to health outcomes. Ahmad et al. showed that the percentage of population living in poverty in communities was associated with a higher cardiovascular and heart failure mortality.^24^ Same investigators have also shown that counties with higher percentage of households living in poor housing conditions had significantly higher risk of COVID-19 incidence and mortality.^14^ In this study, we showed that COVID-19 infection affected communities of distinct income strata in a similar fashion, but with a higher mortality risk in communities of lower household income. Multiple mechanisms have been posited to explain poor health outcomes in low-income population. It is possible that people in lower-income communities have worse health at baseline, receive care at lower quality hospitals, receive differential care within a hospital due to lack of health insurance or poor health literacy, and/or there is a lack of access to care outside of the hospital due to lack of health insurance.^25,26^ We found higher prevalence of obesity, diabetes and smoking in lower income communities to support some of these mechanisms. Moreover, the findings of low-income communities associated with higher COVID-19 mortality risk in non-Medicaid expansion states but no relationship in Medicaid expansion states, strongly suggest that access to health care and health insurance is a potential modifiable risk factor for health disparity in COVID-19 mortality.

Over the past decade, studies have shown that in the states that expanded Medicaid coverage, there were improvements in diagnosis, management and mortality of chronic conditions.^7-10,27-29^ Further studies have also investigated the impact on disease mortality rates in Medicaid expansion states on a nationwide scale.^30,31^ In end-stage renal disease, patients had improved 1-year survival rates in Medicaid expansion states.^31^ Similarly, a decrease in cardiovascular mortality was observed in states after Medicaid expansion. This was considered to be a benefit of improved access to healthcare.^30^ We believe a similar mechanism is at play when it comes to COVID-19 mortality. As shown in our study, in non-Medicaid expansion states, we observed a significant negative association between median household income and COVID-19 mortality which was not seen in Medicaid expansion states. This is likely explained by the improved access to healthcare offered by the ACA which increased access to healthcare insurance of low income individuals by raising the Medicaid eligibility threshold to 138% of the federal poverty level.^32^ A review of literature shows that individuals without health insurance are less likely to seek health care even when in need.^33^ In contrast it has been shown that when they could afford care, individuals were more likely to utilize healthcare resources.^34^ Therefore, while a proportion of population in high income communities are able to afford insurance regardless of state expansion status, we offered evidence that access to health care through Medicaid expansion likely helped close the coverage disparity for low-income individuals in low-income communities from Medicaid expansion states. These results can have policy implications. We posit that improved access to healthcare insurance such as Medicaid expansion could lead to lower uninsured rates among low-income individuals, improving access to healthcare, thereby lowering the adverse impact of COVID-19 in low-income communities.^23,35,36^

### Limitations and Strengths

This is a nationwide study, that utilized representative data of US communities suitable to assess outcomes as it relates to policy. Study limitations include its observational design, inability to conclude causality and the potential for residual confounding despite our careful control of known confounders. Although some states adopted Medicaid expansion into their state constitution during 2020 (Missouri and Oklahoma), none of them achieved implementation stage during 2020.

## Data Availability

All data used in this study were publicly available

## Conclusions and Implications

Median-household-income was not related to COVID-19 incidence but negatively related to COVID-19 mortality in US counties of states without Medicaid-expansion. It was unrelated to mortality in counties of states that adopted Medicaid-expansion. These findings suggest expanded health coverage should be investigated further to attenuate the excessive COVID-19 mortality risk associated with socioeconomically disadvantaged communities.

## ACKNOWLEDGEMENT

The views expressed in this paper represent the authors and not the Department of the Veterans Affairs.

## DISCLOSURES

None

## Notes

### Competing Interest Statement

The authors have declared no competing interest.

### Funding Statement

This work is in part supported by IRP 20-003 (PI: WWC), United States Department of the Veterans Affairs Health Services Research & Development:
https://www.hsrd.research.va.gov/
The funders had no role in study design, data collection and analysis, decision to publish, or preparation of the manuscript.

### Author Declarations

All data used in this study were publicly available; therefore, the study met the criteria for exemption by the Providence Veterans Affairs Medical Center Institutional Review Board.

