## Supplemental Tables 1 and 2 for "“Association between Median Household Income, State Medicaid Expansion Status, and COVID-19 Outcomes Across US Counties”"

Supplement Tables

Table S.1 List of Medicaid Expansion and Non-Medicaid Expansion states as of January 1, 2020

| **Medicaid Expansion States**  **n = 37** | **Medicaid Non-Expansion States**  **n = 14** |
| --- | --- |
| Alaska | Alabama |
| Arizona | Florida |
| Arkansas | Georgia |
| California | Kansas |
| Colorado | Mississippi |
| Connecticut | Missouri |
| Delaware | North Carolina |
| Federal District of Columbia | Oklahoma |
| Hawaii | South Carolina |
| Idaho | South Dakota |
| Illinois | Tennessee |
| Indiana | Texas |
| Iowa | Wisconsin |
| Kentucky | Wyoming |
| Louisiana |  |
| Maine |  |
| Maryland |  |
| Massachusetts |  |
| Michigan |  |
| Minnesota |  |
| Montana |  |
| Nebraska |  |
| Nevada |  |
| New Hampshire |  |
| New Jersey |  |
| New Mexico |  |
| New York |  |
| North Dakota |  |
| Ohio |  |
| Oregon |  |
| Pennsylvania |  |
| Rhode Island |  |
| Utah |  |
| Vermont |  |
| Virginia |  |
| Washington |  |
| West Virginia |  |

| Table S.2 Subgroup analysis of Medicaid and Non-Medicaid SARS-COV-2 mortality as of December 6, 2020 with Median Household Income Quartiles | | | | | | | | |
| --- | --- | --- | --- | --- | --- | --- | --- | --- |
|  | Log Transformed Median Household Income Quartiles | | | | | | | |
|  | **Quartile 1**  Medicaid  N=454  MRR  [95% CI] | **Quartile 2**  Medicaid  N=453  MRR  [95% CI] | **Quartile 3**  Medicaid  N=452  MRR  [95% CI] | **Quartile 4**  Medicaid  N=455  MRR  [95% CI] | **Quartile 1**  Non-Medicaid  N=332  MRR  [95% CI] | **Quartile 2**  Non-Medicaid  N=332  MRR  [95% CI] | **Quartile 3**  Non-Medicaid  N=332  MRR  [95% CI] | **Quartile 4**  Non-Medicaid  N=332  MRR  [95% CI] |
| SARS-COV-2 mortality |  |  |  |  |  |  |  |  |
| (As of December 6, 2020) |  |  |  |  |  |  |  |  |
| Deaths per 100,000 (mean ± SD) | 92.31 ± 128.60 | 71.83 ± 73.10 | 77.18 ± 67.47 | 70.20 ± 138.43 | 138.78 ± 89.11 | 104.13 ± 68.60 | 94.69 ± 79.22 | 73.36 ± 53.55 |
| Model 1 | 1.01 | 1.09 | 0.94 | REF. | 1.43 | 1.31 | 1.15 | REF. |
|  | [0.88-1.15] | [0.97-1.21] | [0.85-1.03] |  | [1.30-1.57] | [1.19-1.43] | [1.05-1.26] |  |
| Model 2 | 1.06 | 1.12 | 0.97 | REF. | 1.41 | 1.28 | 1.14 | REF. |
|  | [0.90-1.26] | [0.98-1.27] | [0.87-1.07] |  | [1.25-1.59] | [1.16-1.42] | [1.03-1.25] |  |
| MRR (95% CI) = Mortality Rate Ratio (95% Confidence Interval)  Model 1: % Population > 65 years, % Male, and % White  Model 2: % Population > 65 years, % Male, % White, Population Density, % Obesity, % Smoking, % Diabetes, Number of Hospitals, Medicaid expansion status according to state policy  *Interaction between income quartiles and Medicaid status was significant (p-value ≤ 0.005) for SARS-COV-2 mortality but not for SARS-CoV2 Cases | | | | | | | | |
